# Two patients with acute meningo-encephalitis concomitant to SARS-CoV-2 infection

**DOI:** 10.1101/2020.04.17.20060251

**Authors:** Raphaël Bernard-Valnet, Beatrice Pizzarotti, Angelica Anichini, Yoris Demars, Enrico Russo, Marie Schmidhauser, Jonathan Cerutti-Sola, Andrea O. Rossetti, Renaud Du Pasquier

## Abstract

Human coronaviruses are known for their tropism for central nervous system and to be associated with neurological complications. SARS-CoV-2 pandemy represent a major health issue and if respiratory symptoms are at the forefront, neurological symptoms should be expected. Here we report the case of two patient that developed encephalitic symptoms with neuropsychological impairment and pathologic cerebrospinal fluid features concomitantly to SARS-CoV-2 documented infection. Both patient recovered promptly without treatment. This report raises the question of central nervous system involvement in SARS-CoV-2 infection and the need for investigation of neuropsychological complications in infected patient.

## Introduction

In December 2019, a cluster of patients with pneumonia of unknown cause led to the identification of a new strain of pandemic coronavirus called Severe acute respiratory syndrome coronavirus 2 (SARS-CoV-2) ^1^. Since the first SARS-CoV outbreak, human coronaviruses are known for their neurological tropism ^2, 3^. If respiratory complications are at the forefront of clinical presentation of SARS-CoV-2, neurological involvement remains poorly described and understood. We report here two patients infected with SARS-CoV-2 who presented with neurological symptoms and signs.

## Methods

Clinical and ancillary tests description was personally retrieved by the authors, who examined the patients. This report is conducted in compliance to Swiss Federal Act on Research involving Human Beings that wave ethic approval for case report of less than 5 patients. Furthermore both patients gave written informed consent for clinical and biological data used for this report. Viral/Bacterial detection were performed using BioFire FilmArray Meningitis/Encephalitis Panel (BioFire Diagnostics, Salt Lake City, USA) and confirmed by traditional polymerase chain reaction (PCR).

### Case descriptions

#### Patient 1

A 64 year-old woman without psychiatric history, known for contact with SARS-CoV-2 (husband tested positive 15 days before) and presenting for 5 days with flu-like symptoms (mild asthenia, myalgia) without fever, acutely developed psychotic symptoms. She was first addressed to a psychiatric ward, but presented a tonico-clonic seizure motivating her admission to an external hospital. A routine electroencephalogram (EEG) revealed nonconvulsive, focal status epilepticus (abundant bursts of anterior low-medium voltage irregular spike-and waves superimposed on an irregularly slowed theta background), managed with intravenous clonazepam and valproate. She was immediately referred to our center. The patient appeared disoriented, with strong attention deficit, verbal and motor perseverations, bilateral grasping, alternating with psychotic symptoms (hyper-religiosity with mystic delusions, visual hallucinations). There was no neck stiffness or focal signs on neurological examination. Cerebral MRI was normal, but her lumbar puncture was compatible with viral meningo-encephalitis (Table 1.), and SARS-CoV-2 was detected in her nasal swab. However, nor SARS-CoV-2 neither classic viral/bacterial pathogens were detected in the cerebro-spinal-fluid (CSF) (Table 1.). Anti-NMDA antibodies were tested negative in CSF. Treatment by acyclovir was transiently administered until herpes simplex/ varicelle zoster virus PCRs came back negative. A follow-up EEG 24h after admission showed a moderate theta background slowing, without epileptiform features. The patient markedly improved 96h after admission with resolution of her symptoms.

**Table 1.**
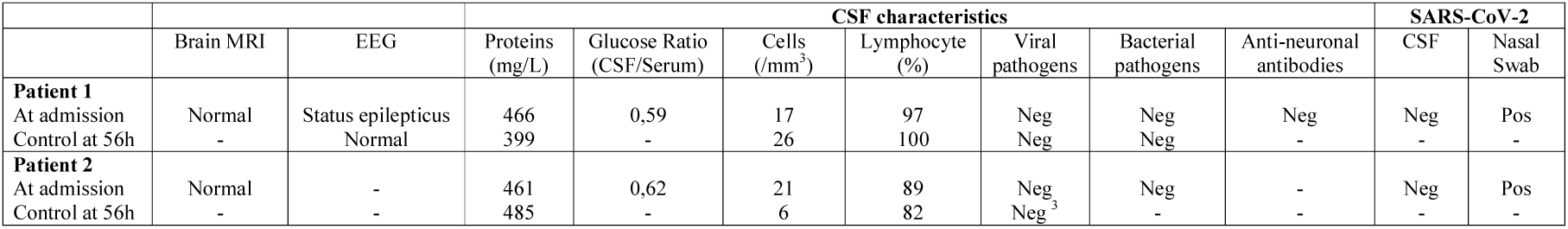
Paraclinal exams in reported patients. Neg: Negative, Pos: Positive. - : not done. Bacterial pathogens: *Neisseria meningitidis, Listeria monocytogenes, Streptoccocus pneumoniae, Haemophilus influenza, Escherichia Coli K1, Streptococcus Agalactiae*; Viral pathogens: Enterovirus, Herpes simplex virus 1, Herpes simplex virus 2, Varicella-Zoster, Cytomegalovirus, Human Herpes Virus 6, Parechovirus. ^3^ Only Herpes simplex virus 1 and 2 PCR were performed. Anti-neuronal antibodies: anti-NMDAR, anti-CAPSR2, anti-LGI1, anti-DPPX, anti- GABA_B_R, anti-AMPAR, anti-lgLON5, anti-mGLUR5, anti-GlyR

|  |  |  | CSF characteristics |  |  |  |  |  |  | SARS-CoV-2 |  |
| --- | --- | --- | --- | --- | --- | --- | --- | --- | --- | --- | --- |
|  | Brain MRI | EEG | Proteins (mg/L) | Glucose Ratio (CSF/Serum) | Cells (/mm<sup>3</sup>) | Lymphocyte (%) | Viral pathogens | Bacterial pathogens | Anti-neuronal antibodies | CSF | Nasal Swab |
| **Patient 1** |  |  |  |  |  |  |  |  |  |  |  |
| At admission | Normal | Status epilepticus | 466 | 0,59 | 17 | 97 | Neg | Neg | Neg | Neg | Pos |
| Control at 56h | - | Normal | 399 | - | 26 | 100 | Neg | Neg | - | - | - |
| **Patient 2** |  |  |  |  |  |  |  |  |  |  |  |
| At admission | Normal | - | 461 | 0,62 | 21 | 89 | Neg | Neg | - | Neg | Pos |
| Control at 56h | - | - | 485 | - | 6 | 82 | Neg<sup>3</sup> | - | - | - | - |

#### Patient 2

A 67 year-old woman, already diagnosed for SARS-CoV-2 infection for 17 days with mild respiratory symptoms, presented an intense wake-up headache. Few hours later, she was found drowsy and confused, lying on the ground of her bathroom. She was referred to our hospital. On neurological evaluation, she was disoriented with motor perseverations, bilateral grasping, aggressiveness and left hemianopia and sensory hemineglect; there was no neck stiffness. A SARS-CoV-2 pneumonia was diagnosed thanks to a positive nasal swab and an ultrasound showing subpleural condensation. Brain MRI was normal and her lumbar puncture revealed lymphocytic pleocytosis (Table 1). Yet, CSF SARS-CoV-2 and viral/bacterial pathogens PCR were negative (Table 1). The patient transiently received ceftriaxone, amoxicillin and acyclovir. Neurological symptoms resolved within 24 hrs, except for a mild headache. The patient was discharged 72 hrs after admission with no symptoms.

## Discussion

We report on two patients who developed a meningo-encephalitis few days after a diagnosis of SARS-CoV-2 infection. Both had “benign” form with only mild respiratory and general symptoms. Yet, they suddenly developed severe neuropsychological symptoms and a status epilepticus for one of them. The CSF profiles being compatible with viral meningo-encephalitis, a large screening for usual pathogens, including SARS-CoV-2, was performed but came back negative. While the proof of a direct involvement of SARS-CoV-2 is missing, we hypothesize that the latter virus was responsible for this neurological presentation. First, the usual suspects causing viral meningo-encephalitis were tested negative. Second, the neurological picture occurred in the wake of proven SARS-CoV2 infection. Third, coronaviruses are known for their neurological tropism and for inducing encephalitis. Of note CSF detection of coronavirus RNA seems infrequent ^3^. A possible mechanism accounting for the encephalitic presentation in these patients may be a para-infectious one, somewhat reminiscent of the coronaviruses association with acute disseminated encephalomyelitis and (for SARS-CoV-2) Guillain-Barré syndrome ^4, 5^. Such mechanism would explain the rapid clinical recovery of both patients and the absence of MRI lesions, suggesting a limited viral process, contrary to a previous reports showing severe encephalitis and viral RNA in the CSF, although, in this case, herpes simplex virus encephalitis was not formally excluded ^6^.

To conclude, we report the first temporal association between acute SARS-CoV-2 infection and aseptic encephalitis with focal neurological symptoms and signs. Further studies are needed to identify the spectrum of neurological complications of this pandemic outbreak and the underlying pathophysiological mechanisms.

## Data Availability

-

## Aknowledgement

We would like to thanks Prof. Pierre-Alexandre Bart, Dr. David Gachoud, Dr. Jan Novy, Dr. Nicola Marchi and Dr. Sergiu Vijala for taking care of these patients at different steps during their hospitalization. We also would like to acknowledge the work of Dr. Onya Opota, Dr. Katia Jaton and Prof. Gilbert Greub for molecular biology diagnostic testing.

## Appendix 1 Authors

Appendix 1 - Authors
| **Name** | **Location** | **Contribution** |
| --- | --- | --- |
| Raphaël Bernard-Valnet, MD, PhD | Centre Hospitalier Universitaire Vaudois, Lausanne, CH | Design and conceptualized study; analyzed the data; drafted the manuscript for intellectual content |
| Beatrice Pizzarotti, MD | Centre Hospitalier Universitaire Vaudois, Lausanne, CH | Collected the data; drafted the manuscript for intellectual content |
| Angelica Anichini, MD | Centre Hospitalier Universitaire Vaudois, Lausanne, CH | Collected the data |
| Yoris Demars, MD | Centre Hospitalier Universitaire Vaudois, Lausanne, CH | Collected the data |
| Enrico Russo, MD | Centre Hospitalier Universitaire Vaudois, Lausanne, CH | Collected the data |
| Marie Schmidhauser, MD | Centre Hospitalier Universitaire Vaudois, Lausanne, CH | Collected the data |
| Jonathan Cerutti-Sola, MD | Centre Hospitalier Universitaire Vaudois, Lausanne, CH | Collected the data |
| Andrea O. Rossetti, MD, FAES | Centre Hospitalier Universitaire Vaudois, Lausanne, CH | Revised the manuscript for intellectual content |
| Renaud Du Pasquier, MD | Centre Hospitalier Universitaire Vaudois, Lausanne, CH | Design and conceptualized study; revised the manuscript for intellectual content |

